# Depression is a more significant predictor for wellbeing in Inclusion Body Myositis than physical disability

**DOI:** 10.1101/2024.01.22.24301628

**Authors:** G. Nunn, G. Glenister, K. Hird, K. Beer, I. Cooper, M. Needham, A. Brusch

## Abstract

**Objectives:** (1) determine if there is a correlation between disability, depression, and wellbeing in people with IBM, (2) determine if disability and depression can predict wellbeing in people with IBM, and (3) identify the prevalence of depression and impaired wellbeing in participants with IBM.

**Methods:** In this cross-sectional study, 101 participants with IBM completed the Neuromuscular Symptom Score (NSS), Personal Wellbeing Index (PWI), and Patient Health Questionnaire-9 (PHQ-9) surveys to serve as surrogate measures of physical disability, wellbeing, and depression respectively.

**Results:** Linear regression identified that PHQ-9 significantly predicts PWI, however NSS does not, with a negative predictive value of depression for wellbeing (−2.7513, p < 0.001) and a positive predictive value of disability for wellbeing (0.0575, p = 0.764). Moderate to severe depression was reported in 78.2% of participants, and all but one participant reported reduced wellbeing.

**Conclusions:** Depression is a more significant predictor of wellbeing than disability in participants diagnosed with IBM. There was a high prevalence of depression and reduced wellbeing in participants, highlighting the importance of assessing these factors to optimise treatment in IBM.

## Background

When a person with a progressive neurological disease presents to a clinician, it is easy to assume that the more debilitating the disease, the worse the person’s quality of life, wellbeing, and mental health. However, anecdotally this does not seem to be the case. Many patients with severe disability limiting their function still report high wellbeing and good quality of life. Conversely, patients with comparatively little disability report a significant impact on their wellbeing and mental health.

IBM is a rare and progressive degenerative disease affecting skeletal muscle. It is characterised by a gradual decline in muscle strength, leading to considerable impairment in everyday activities such as walking, fine motor skills, and swallowing^1^. Unlike other forms of myositis, IBM does not typically respond to immunosuppressive treatments, thereby posing a significant challenge in the management of the disease^2^. Current IBM treatment is limited to physiotherapy and exercise programs^3^. Consequently, IBM patients often experience a protracted period of progressive functional decline spanning decades post-diagnosis. Given this observed discrepancy this project was designed to explore the relationship between wellbeing and disability associated with inclusion body myositis (IBM) and to quantify the prevalence of depression in IBM participants.

Where health-related quality of life involves a cognitive assessment an individual makes regarding the impact of their disease, wellbeing reflects their emotional response^4^. Wellbeing is more closely linked to mental health and is a state of being that enables a person to thrive^5^. Multiple studies have shown that positive wellbeing leads to better physical health including better immune function, though this has not specifically been studied in autoimmune disease^6-8^.

A person does not have to be without negative emotions to have sustained positive wellbeing; it is the presence of long term or extreme negative emotions that have a detrimental effect^9^. Many factors are known to contribute to wellbeing including personality (e.g extraversion is associated with positive wellbeing)^10^, demographics (young or elderly is associated with positive wellbeing)^11^, and socioeconomic factors (reduced income inequality^12^ and being employed are associated with positive wellbeing^13^). These factors are complex and not easily modifiable, if at all.

Positive mental health as defined by the World Health Organisation is when a person can realise their abilities, can cope with the normal stresses of life, can work productively, and are able to contribute to their community^14^. Disability and morbidity secondary to IBM can interfere with these factors thereby impacting on mental health.

This interplay between wellbeing, mental health, and disability due to chronic disease is the basis for the argument health-related quality of life can be improved if wellbeing and mental health are actively treated. Hence, it is important to consider a person’s wellbeing and mental health as an integral aspect of the clinical assessment. The Neuromuscular Symptom Score (NSS) survey, the Personal Wellbeing Index (PWI), and the Patient Health Questionnaire (PHQ-9) are three measures that have been used to capture patient perspectives of disease severity and burden.

The NSS survey is designed to quantify participant appraisal of their disease severity. The survey, which is not disease specific, has been used in multiple studies to assess all forms of myositis^15-17^ and comprises two parts: Part A assesses fine and gross motor function through everyday activities, and consists of fifteen questions. Questions are scored using a 4-point scale ranging from 0-3 where 0 = total dependence and 3 = normal function. Part B evaluates fatigue and tripping using 5 questions where 0 = always and 3 = never. By self-reporting their ability to perform these activities, participants provide a surrogate measure of the disease’s impact on their lives.

The PWI, validated by Khor et al., in 2013 and recommended by the World Health Organization (WHO) and the Organisation for Economic Co-operation and Development (OECD), assesses self-reported wellbeing across seven domains: standard of living, health, achieving in life, relationships, safety, community connectedness, and future security^18^. Total scores are categorized into “normal levels of subjective wellbeing” (score of 70+), “compromised level of subjective wellbeing” (50-69), or “challenged level of subjective wellbeing” (<50).

The PHQ-9, validated by Kroenke et al., serves as a measure of depression severity and diagnostic tool^19^. Comprising nine questions that assess symptoms over a two-week period, total scores can be interpreted as minimal depression (score of 1-4), mild depression (5-9), moderate depression (10-14), moderately severe depression (15-19), or severe depression (20-27).

Previous studies have focused on quality of life in IBM. Sadjadi et al. found that functional disability did result in reduced quality of life however mood had a more direct effect^20^. It is noteworthy that quality of life can paradoxically be higher in more severe, longer-lasting diseases due to a “response shift to diagnosis”, whereby individuals acclimate to their altered health circumstances^21^. In the case of IBM, where treatment options are limited primarily to physical therapy, patients may adapt to their condition over time. While previous research has reported reduced quality of life in myositis participants with higher rates of disease impairment measured by muscle strength, daily activity performance, and symptoms of fatigue and pain^22-23^, there is a dearth of studies exploring wellbeing and depression in IBM. A clearer measure of association between IBM disability, wellbeing, and depression will support the notion that wellbeing and depression should become an integral aspect of assessment and treatment in IBM.

The aim of this study is to identify if disability or mental health are a stronger predictor for wellbeing in participants with IBM. More specifically, the aims are (1) determine if there is a correlation between disability, depression, and wellbeing in people with IBM, (2) determine if disability and depression can predict wellbeing in people with IBM, and (3) identify the prevalence of depression and impaired wellbeing in participants with IBM. It is hypothesised that depression will be more strongly correlated with wellbeing than disability. This study is important to optimise care delivered to people with IBM by highlighting the non-physical aspects of the disease often overlooked.

## Methods

### Recruitment

This study was a cross-sectional exploration of wellbeing and the presence of depression in people with IBM in Australia. It involved analysis of data from participants who were surveyed as part of a larger study on incontinence in myositis. The original study included the three surveys used in this study, as well as an original survey to assess the prevalence of incontinence. This study had ethical approval from the Murdoch University Human Research Ethics Committee, with participants giving informed consent prior to commencing the surveys.

A customised survey using Qualtrics^®^ was distributed to an estimated 300 people via the Myositis Association Australia and the Myositis Discovery Programme clinics in Western Australia. There were potentially participants contacted through both groups (estimated 50 people overlap). Due to study anonymity, we were unable to determine exact numbers. To avoid repeat data entry, Qualtrics^®^ only accepted one response per internet protocol (IP) address. Response rate was 59.6%. The survey was open for 8 weeks.

To be included in the study, participants had to have a confirmed diagnosis of inclusion body myositis by a neurologist.

### Patient and Public Involvement

This study evolved from the observation of paradoxically better wellbeing reported in patients with more physically disabling IBM. Through discussions with patients, it was noted that many reported symptoms of depression as having a big impact on their life. A consumer engagement panel was also informed of the study to disseminate the survey and study results to the members of the Myositis Association Australia and Myositis Discovery Programme Clinics.

### Surveys

Participants completed the Neuromuscular Symptom Score survey, the Personal Wellbeing Index, and the Patient Health Questionnaire-9. The outcome of each survey response is reported as a total score that serves as a surrogate measure of overall participant reported disability, wellbeing, and severity of depression respectively.

## Results

In the original recruitment for the continence survey, 149 participants responded, of which 101 had inclusion body myositis. Of these, 95 participants completed the NSS, PWI, and PHQ-9, with a further 6 participants completing only the NSS and PHQ-9.

### Statistical Analysis

**Table 1:** Cohort demographics.

| Variable |  | Frequency (%) |
| --- | --- | --- |
| Sex | Female | 40 (39.6) |
|  | Male | 61 (60.4) |
| Age Range | 40-49 | 1 (1.0) |
|  | 50-59 | 4 (4.0) |
|  | 60-69 | 23 (22.8) |
|  | 70-79 | 57 (56.4) |
|  | 80-89 | 15 (14.9) |
|  | 90+ | 1 (1.0) |

To examine wellbeing in participants, scores were divided into each PWI category (normal, compromised, and challenged level of subjective wellbeing) as per the survey’s validated categories^18^.

**Figure 1:**
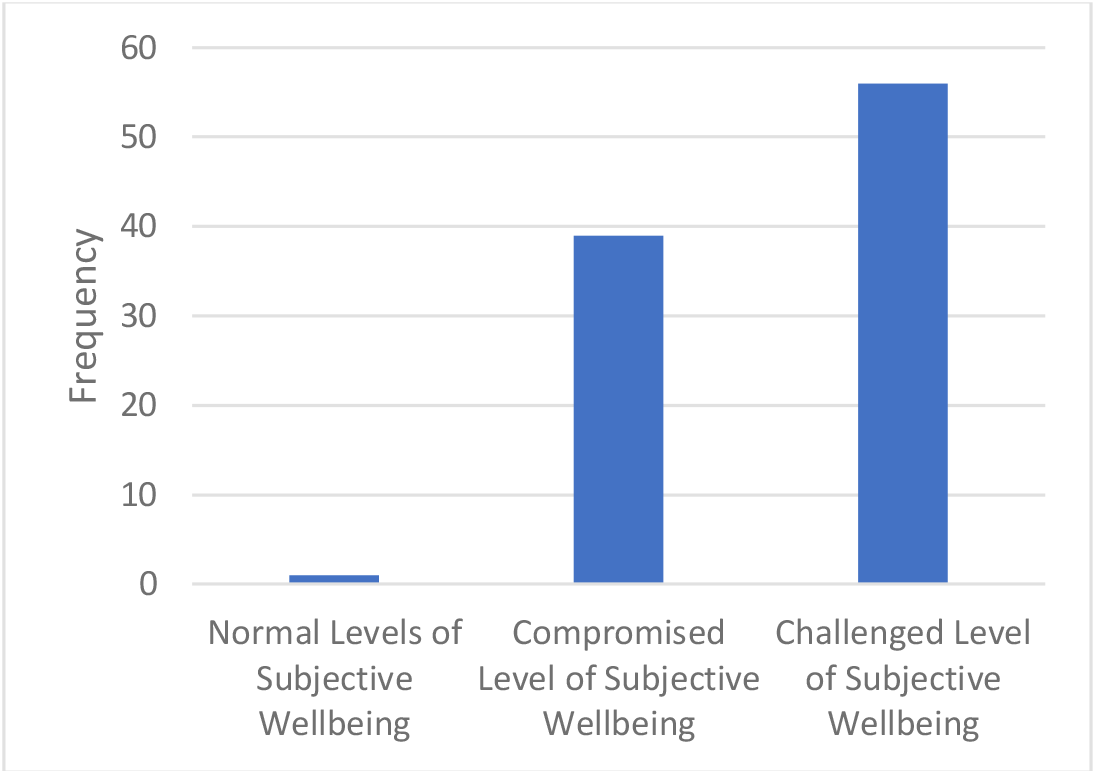
Frequency of wellbeing scores per category as assessed by PWI.

To examine the prevalence of depression, participants were divided into each PHQ-9 category (minimal, mild, moderate, moderately severe, and severe depression) as per the survey’s validated categories^19^.

**Figure 2:**
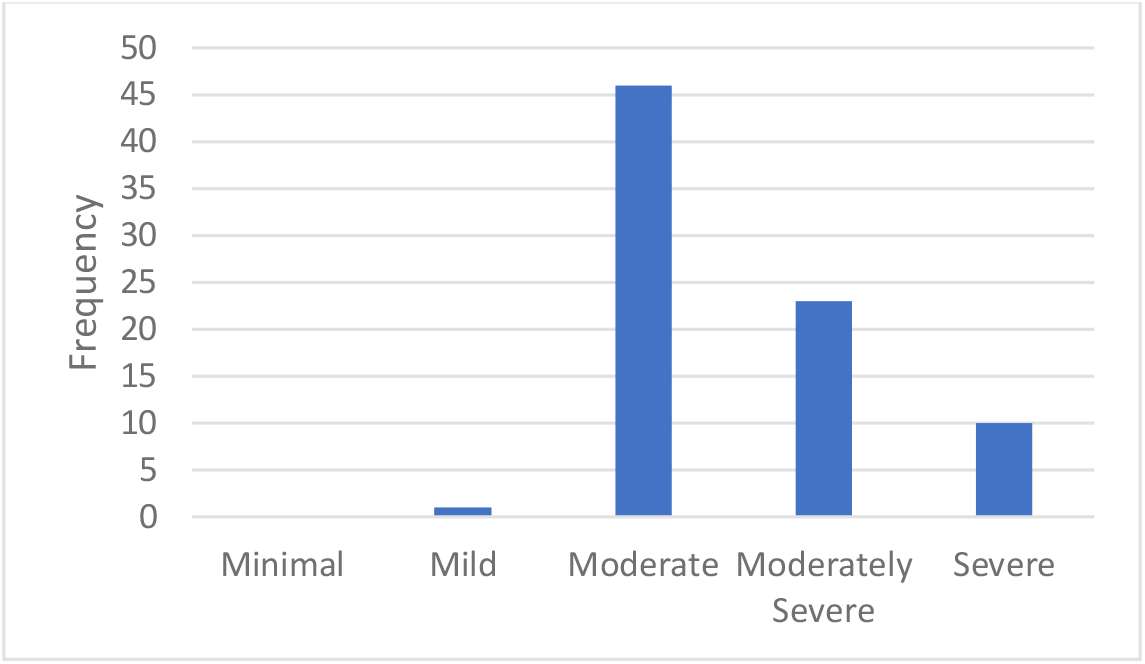
Frequency of depression severity scores per category as assessed by PHQ-9.

PWI scores were correlated with NSS scores to determine if there was a relationship between wellbeing and disability. Higher PWI scores indicate positive wellbeing, and higher NSS scores indicate less disability.

**Figure 3:**
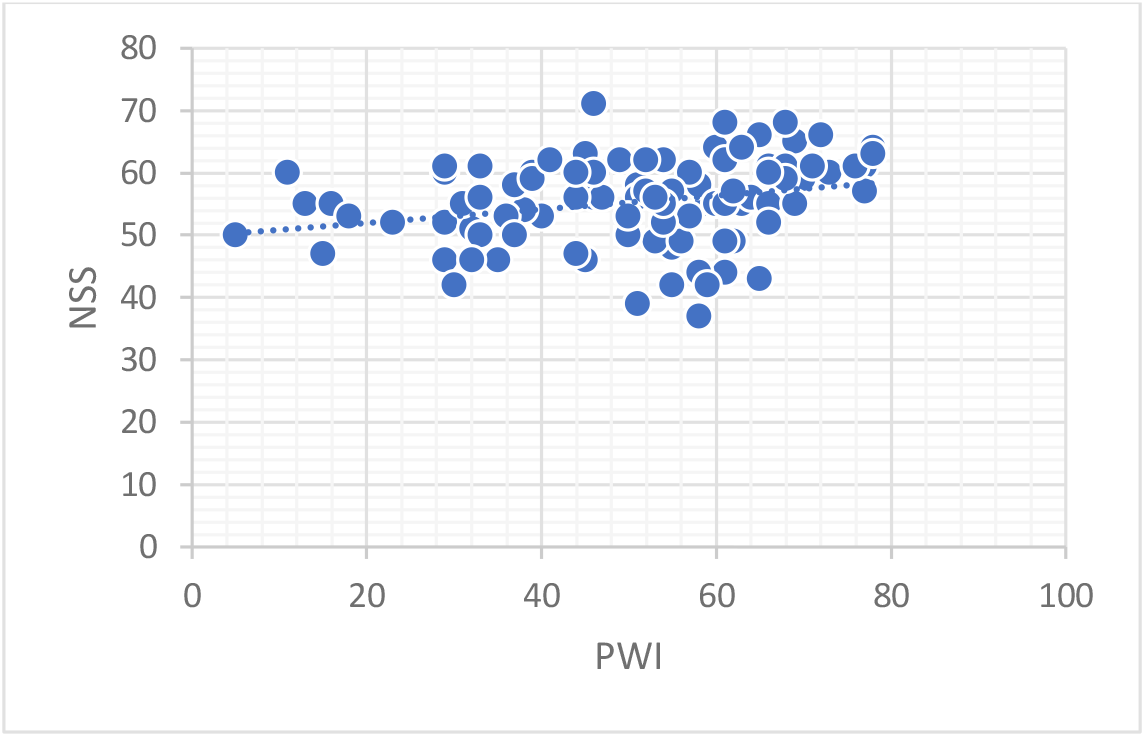
Correlation scatterplot of disability (NSS) and wellbeing (PWI), Pearson’s r value 0.265 (p-value 0.009)

PHQ-9 scores were then correlated with NSS scores to determine if there was a relationship between depression and disability. Higher PHQ-9 scores indicate more severe depression, and higher NSS scores indicate less disability.

**Figure 4:**
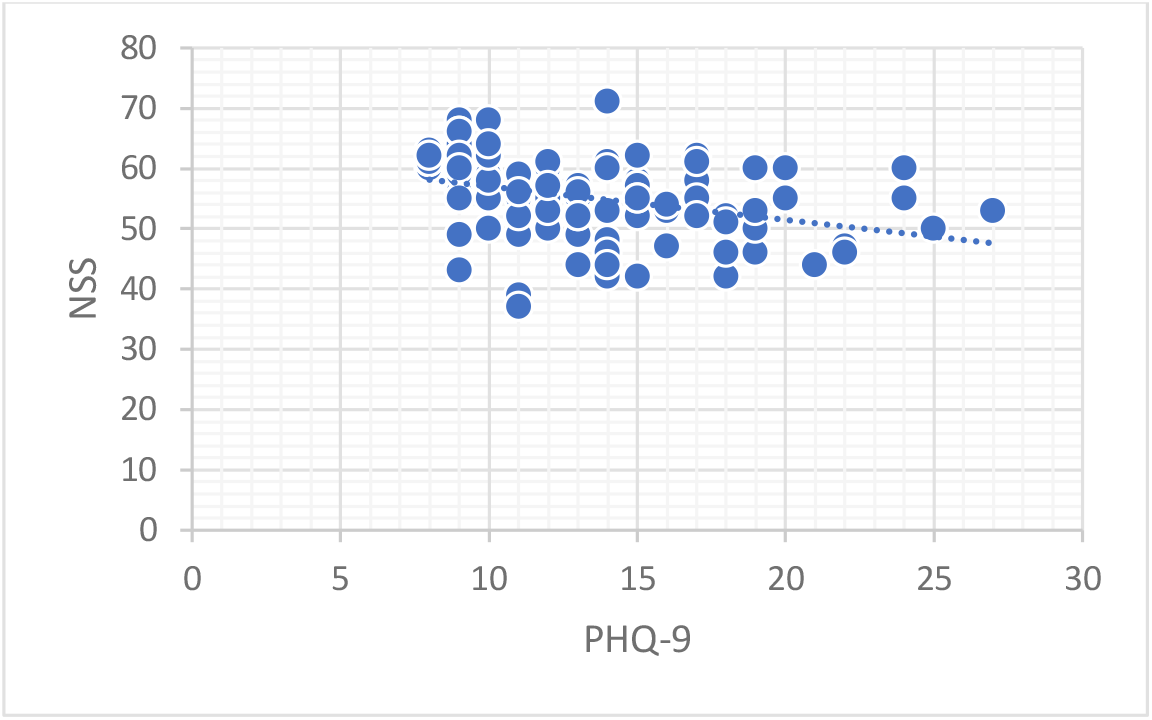
Correlation scatterplot of disability (NSS) and depression (PHQ-9), Pearson’s r value -0.353 (p-value <0.001)

Finally, PWI scores were correlated with PHQ-9 scores to determine if there was a relationship between wellbeing and depression. Higher PWI scores indicate positive wellbeing, and PHQ-9 scores indicate more severe depression.

**Figure 5:**
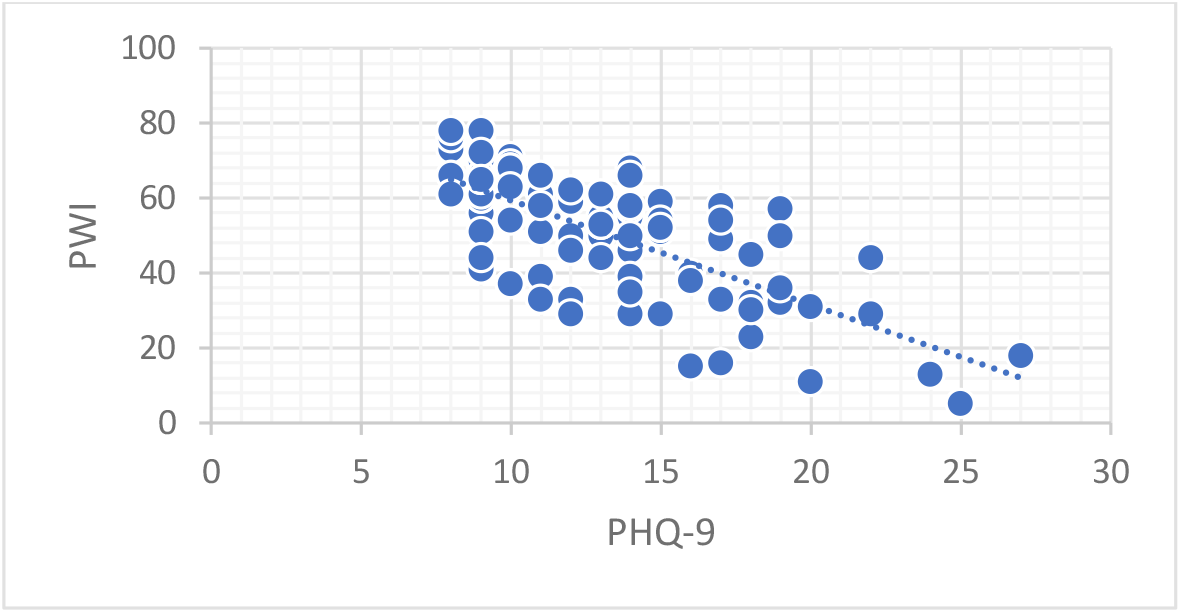
Correlation scatterplot of wellbeing (PWI) and depression (PHQ-9), Pearson’s r value -0.702 (p-value <0.001)

Simple linear regression tested whether disability (NSS) and depression (PHQ-9) predict wellbeing (PWI). The model generated was significant, with a p-value of <0.001 and accounts for a variance of 49.4% (r^2^ = 0.494). It was found that PHQ-9 significantly predicts PWI, however NSS does not.

**Table 2:** Linear regression values for predicting PWI using PHQ-9 and NSS.

| Predictor | Estimate | SE | t | p |
| --- | --- | --- | --- | --- |
| Intercept | 83.5953 | 12.683 | 6.591 | <0.001 |
| PHQ-9 | -2.7513 | 0.314 | -8.771 | <0.001 |
| NSS | 0.0575 | 0.191 | 0.301 | 0.764 |

## Discussion

The hypothesis that depression is a stronger predictor for wellbeing in participants with IBM than disability was supported by our results. Wellbeing and depression were highly varied in IBM participants regardless of degree of disability, as seen by the overall model fit. These results support our argument that wellbeing cannot be assumed in patients with IBM and that there is a role for assessing patient wellbeing and mental health, as well as their physical and functional disability.

Wellbeing (PWI survey) showed a significant positive correlation with participant’s appraisal of their disability revealing that better wellbeing was seen in participants with less disability. However, it had the lowest correlation value of the three surveys compared. Therefore, we can infer that disability does contribute to wellbeing, but plays a less important role than depression.

Wellbeing is reduced overall among IBM participants, as seen when scores are split into the categories “normal levels of subjective wellbeing”, “compromised level of subjective wellbeing” and “challenged level of subjective wellbeing”, with only one participant scoring a normal level of subjective wellbeing. While we cannot directly compare these results to the Australian population who do not have IBM, it does paint a convincing picture that wellbeing is significantly impaired.

To the best of our knowledge, this is the first study to thoroughly assess wellbeing in IBM. Similarly to our study, Armadans-Tremolosa et al. found no association between wellbeing and the physical symptoms of dermatomyositis and polymyositis, concluding that psychological wellness is likely independent from disease severity^22^. Multiple studies support the notion that quality of life in myositis is affected by disease severity and mood, but wellbeing has not been sufficiently explored in its own right^24-27^.

Depression (PHQ-9) was negatively correlated with disability, where more severe depression was seen in participants with greater disability. This is a stronger correlation with disability than wellbeing. The results of this study revealed symptoms of moderate to severe depression in 78.2% of participants. This finding contrasts with a study by Senn et al., who found only 37.8% of their participants had symptoms of depression at all^28^. The study assessed participants with IBM in Germany via the Hospital Anxiety and Depression Scale German version, therefore a direct comparison is not possible. Varied prevalence of depression could also be due to alternative cultural and societal factors. Both studies show a much higher prevalence of depression than expected given prior study findings. Moussavi et al. explored data from 60 countries and found that there was between 9.3% to 23.0% prevalence of depression in participants with a comorbid chronic physical disease (though none of the participants had IBM)^29^. This prevalence was also higher than participants who did not have a comorbid physical disease (3.2% 1-year prevalence)^29^.

Wellbeing and depression were most strongly correlated to each other, as expected given that wellbeing is known to be strongly tied to mental health^5^. The results of this study support this association as scores for mental health were a stronger predictor of wellbeing than physical disability.

## Limitations

The findings of this study need to be considered in the context of the following limitations. Firstly, the NSS, where we attributed disability to IBM, cannot exclude contributions from other diseases that impact a person’s ability to perform various tasks. Secondly, the variation in the surveys used across different studies as surrogate measures for disability, wellbeing, and depression complicates direct comparisons with existing research. Thirdly, the recruitment method, relying on email outreach to willing survey participants, may introduce selection bias to participants more likely to engage with services (especially as treatment for IBM is physical therapy), potentially skewing the results towards better wellbeing or reduced disability. Given that the initial study focused on continence there may have been a response bias in that only people reporting continence issues completed the survey. Lastly, the study has a relatively small sample size of 101 participants and the results revealed considerable variability. To be more confident in the results, replicating the study with a larger sample size would be ideal.

Future research could focus on finding and validating the best tools to assess depression and wellbeing in IBM patients in a clinical setting. It should also be assessed whether subjective and objective neuromuscular assessments differ in their correlation to wellbeing and depression, given the choice to use self-reported disability where clinical assessments rely more on specialist assessment. Other mental illnesses in people with IBM, such as anxiety disorders, could be explored as our study and many others focus only on depression. Finally, factors contributing to reduced wellbeing in patients with IBM could be explored further to create a path towards more personalised treatment.

## Conclusion

This study shows that self-reported scores on a depression scale is a more significant predictor of wellbeing than disability in participants diagnosed with IBM. There was a high prevalence of depression in the participants, even accounting for a comorbid physical condition. Given the interplay between wellbeing, depression, and disability, incorporating a focus on wellbeing into treatment may be a path towards improving health-related quality of life in IBM patients.

## Data Availability

All data produced in the present study are available upon reasonable request to the authors

## Key points

- Depression is a greater predictor for wellbeing than disability in inclusion body myositis.
- Wellbeing was significantly reduced and depression symptoms severe in participants with inclusion body myositis.
- Clinicians should include regular assessments on wellbeing and mental health to significantly improve quality of life in people with inclusion body myositis.

## Notes

### Competing Interest Statement

The authors have declared no competing interest.

### Funding Statement

This study did not receive any funding

### Author Declarations

HREC of Murdoch University gave ethical approval for this work

